# COMORBIDITY ANALYSIS: OVERLAPPING SEMICIRCLES WITH WIGNER LAW AND RANDOM MATRIX THEORY

**DOI:** 10.1101/2021.08.23.21262184

**Authors:** O. Nolasco-Jáuregui, L. A. Quezada-Téllez, Y. Salazar-Flores, Adán Díaz-Hernández

## Abstract

In December 2019 COVID-19 appeared as a new pandemic that has claimed the lives of millions of people around the world. This article presents a regional analysis of COVID-19 in Mexico. Due to the comorbidities of Mexican society, the new pandemic implies a higher risk for the population. The study period runs from April 12 to October 5, 2020 (761 665 Patients). In this proposal we apply a unique methodology of random matrix theory in the moments of a probability measure that appears as the limit of the empirical spectral distribution by the Wigner semicircle law. The graphical presentation of the results is done with Machine Learning methods in the SuperHeat maps. With this is possible to analyze the behavior of patients who tested positive for COVID-19 and their comorbidities. We conclude that the most sensitive comorbidities in hospitalized patients are the following three: COPD, Other Diseases and Renal Diseases.

## 1. Introduction

Throughout its history, humanity faced different pandemics were millions of people lost their lives in the world. The recently epidemic of those SARS-CoV and MERS-CoV stand out [1]. Currently in December 2019 in the city of Wuhan-China, a series of cases were reported that met criteria for pneumonia with severe characteristics. Due to them, the local health authorities noticed that in the patients an epidemiological relationship with a wholesale seafood market, where wild animals were also sold [2].

For December 31 it was notified to the Chinese Center for Disease Control and Prevention an epidemiological investigation, like at first security measure was the closing of the seafood market to the public on January 1, 2020. Later on January 9, the Chinese government reported the discovery of the new coronavirus; and on January 12 they released their genomic sequence of nCoV-2019. Initially, the epidemic growth rate was 0.10 per day (95% CI) and it was doubling time in 7.4 days. On January 11, the first death was reported in China [1].

On January 13 in Thailand, the first imported case was registered in a 61-year-old patient from Wuhan. The USA reported its first confirmed case on January 20 in a 35-year-old patient who traveled to Wuhan. It was until January 30 that the WHO declared the nCoV-2019 infection an international public health emergency. On February 11, the name of the disease officially changed to COVID-19 (coronavirus disease). The name of the virus, after genomic analysis of the sequences, is SARS-CoV-2 [3].

COVID-19 arrives in Mexico in February 2020. On February 27, 2020, it was announced in the media that one patient had tested positive to the virus. This patient who tested positive went to the INER, where he mentioned having traveled to Bergamo Italy; there, he had contact with an infected person. On February 28, the Institute for Diagnosis and Epidemiological Reference to “Dr. Manuel Martínez-Báez” (InDRE) confirmed the first case of COVID-19 in Mexico. Following up four more cases, they found that they traveled to Italy too; which three of them had mild symptoms. Two of these patients stayed in Mexico City and one in Sinaloa. The fourth patient did not develop symptoms, so he was a carrier. Perhaps this was the first reported asymptomatic patient in Mexico. The following days new cases were presented, as a result on March 1st, all cases in Mexico were imported [2].

Random Matix Theories (RMTs) have grown enormously in fields such as wireless communication theories [4], biology in RNA analysis [5], pure mathematics [6], probability [7], among others [8]. The RMTs [8] are a set of matrices in real symmetric bands with inputs extracted from an infinite sequence of interchangeable random variables, as far as the symmetry of the matrices allows it [9]. The entries of the upper triangular matrices of are correlated and these correlations are not assumed to be small or sparse [10]. The RMTs have in their eigenvalues distribution measures still converge in a semicircle but with a random scale [11], also they have asymptotic behavior of the norm attributions in 2 operators [12]. The key to his analysis is a generalization of a classical Finetti result that allows the underlying probability spaces to be represented as averages of the Wigner band sets with the inputs not necessarily centered [13]. Some results appear to be new even for such Wigner band matrices [14].

Wigner has been used with operators in large data dimensions with independent input the Random Matrix Theory (RMT) [14]. Wigner has also analyzed the distribution of the gaps in the energy levels, where they found that they were independent of the underlying matter; surprisingly, this gap distribution is successfully reproduced by RMT [15].

This study focuses on a particular case in Mexico and it is undoubtedly that this methodology is applicable to many countries in the world. One of the approaches proposed in this research is through the Random Matrix Theory (RMT) approach. RMT has its origin and application when John Wishart analyzed properties in multivariate normal populations [16]. Also in the predictions in quantum mechanics, the energy levels can be able calculated by the eigenvalues together with the RMT elements [17]. In general the RMTs can work to analyze the multivariate behavior of data as it is done in this work. One of the most important contributions of this document is Wigner and RMT in comorbidities of patients with COVID-19.

The effect of the COVID-19 pandemic in Mexico has been investigated by some authors, like [18], they used data mining for data analysis. In [19] they analyzed the risk factors for COVID-19 and managed to rank the most determining factors using a multivariate logistic regression. In [20] in 2021 they found a high incidence of comorbidities in deaths that occurred up to August 2020. In and another analysis, [21] in 2021 give predictions on the spread of the pandemic using Bayesian inference.

However, as far as we know, there is no study that uses our methodology that has analyzed the Mexican case. This methodology describes a tool which helps to infer weak convergence: with the method of moments in a probability measure. We propose to apply this method for both deterministic and random probability measures. This document studies in depth the concepts of weak convergence of probability measures and random probability measures with the Wigner law.

In this application it is important to know the moments of a probability measure or at least some properties of the moments in combination with the fact that a real symmetric matrix is positive definite in the real sense. Oftentimes it will not be of interest if a sequence of numbers really belongs to a probability measure, since we automatically obtain this result when employing the method of moments. This method a priori assuming that the target distribution to have specific moments, so, it can be used to check convergence to a random probability measure. In any case, the essential for the method of moments is the knowledge about the uniqueness of a distribution with given moments, that is, there is at most one distribution with a given sequence of moments. In random matrix theory, the probability measure that appears as the limit of the empirical spectral distribution is a naturally of the semicircle distribution. What we mean by naturally? is that it appears in Wigner’s semicircle law, which is the easiest non-trivial random matrix ensemble, for it has standardized entries which are independent up to the symmetry constraint. It is safe to say that the role of the semicircle distribution in random matrix theory is as large as the role of the standard normal distribution in probability theory. To remind the reader, the semicircle distribution is the probability measure.

## 2. Method

### 2.1 Definitions of the Theory of Random Matrices

The Wigner matrices are unit matrices, written in an irreducible unit group (SU) and their rotationally (SO) matrices [11]:

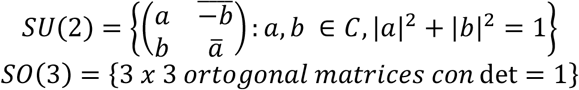

where Jx, Jy, and Jz are generators of the *Lie algebra* of the previous groups [22], that is, there is a non-associative vector of space g, with an alternate bilinear map: g x g ⇥g; (x, y) ⇥ [x, y], satisfying the Jacobi identity, which means that the sum of all even permutations is zero. So these three operators are the components of a vector operator, known as angular momentum.

#### Definition 1. Hermitiana matrix

A square matrix *A* ∈ *M*_*n*_ (*C*) called Hermitian matrix if it has the property of A * = A, where A * denotes the conjugate transpose (or Hermitian transpose) of A, that is, where the subscripts _i, j_ are formally defined by 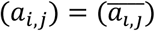. An important property of these matrices is that each Hermitian matrix is diagonalizable and its eigenvalues are real and its eigenvectors are two by two orthogonal [11].

#### Definition 2. Probability Density Function

The probability density function f_x_(t) of a continuous random variable, is a function whose value at any given sample (or point) in the sample space (the set of possible values taken by the random variable) can be interpreted as providing a relative likelihood that the value of the random variable would equal that sample. A distribution has a density function if and only if its cumulative distribution function F_X_(x) is absolutely continuous. In this case: F is almost everywhere differentiable, and its derivative can be used as probability density [13]:

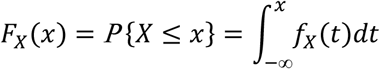

#### Definition 3. The empirical measure

If X_1_, X_2_, … be a sequence of identically distributed independent random variables with values in R. Where is denoted by P their probability distribution. The empirical measure of P_n_ is a measurable subset A ⊂ R. The empirical distribution function is an estimate of the cumulative distribution function that generated the points in the sample. It converges with probability 1 to that underlying distribution, according to the Glivenko–Cantelli theorem. A number of results exist to quantify the rate of convergence of the empirical distribution function to the underlying cumulative distribution function [13]:

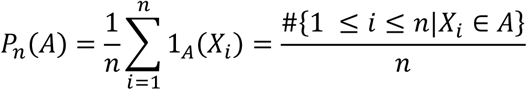

where 1A is the indicator function. Please note that if it chooses A =[-∞, *x*], ∀*x* ∈ *R*, then P_n_ (A) is the distribution of the empirical function.

#### Definition 4.

A Wigner matrix W_n_ ∈ M_n_ (C) is a Hermitian matrix where (X_i, j_) with subscripts i <j such that X_i, j_ are independent and identically randomly distributed variables and are of the complex type with i <j [14]:

- Xi, j are independent and identically distributed real random variables
- E [Xi, j] = 0, ∀i, j
- *E* [[*X*_*i, j*_]^2^] = *s*^2^, *if i* ≠ *j*
- *E* [[*X*_*i, j*_]^2^] < ∞

#### Remark 1.

Some iconic Wigner sets are the Gaussian Unitary Ensemble GUE (n) and the Gaussian Orthogonal Ensemble GOE (n), are described by the Gaussian ensembles measure density. GUE Consider a complex Wigner matrix where Xi,j is standard complex Gaussian (i.e. Xi,j ∼ N(0, 1 2) + iN(0, 1 2)) and Xi,i ∼ N(0, 1) (real), which are defined as follows [9]:

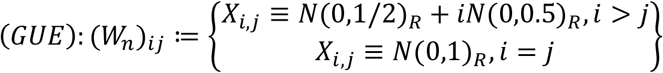

Let C ∈ C^nxn^ unitary, then CC^*^ = I and C^*^W_n_C has the same distribution as W_n_, that is (GOE) is invariant under unit conjugation. GOE is real Wigner matrix where Xi,j ∼ N(0, 1) and Xi,i ∼ √ 2N(0, 1) [23]:

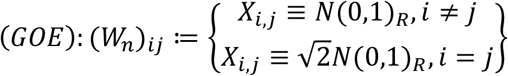

Where C ∈ R^nxn^ orthogonal, then CC^T^ = I and C^T^W_n_ C has the same distribution as W_n_, that is, GOE(n) is invariant function under orthogonal conjugation. Now, the focus is on Gaussian Wigner matrices, whose inputs are Gaussian random variables with zero mean and variance s^2^ if i ≠ j and 2s^2^ if i = j, but this theory only applies to general distributions [23].

#### Definition 5. The Operator Norm for Band Random Matrices

The semicircle law for Wigner band ensembles suggests that in the case of centered entries the operator norm should asymptotically be of the order of the square root of the bandwidth. It was already observed in [23-24] that this cannot hold if the bandwidths do not grow at least at some logarithmic rate with the matrix size.

In the first subsection, we provide in Definitions (1-4) and in Remark 1 positive results in this direction that guarantee for centered Wigner band ensembles an almost sure upper bound on the operator norm that grows proportionally and the bandwidth satisfies some growth condition. The second subsection considers the situation of Wigner band ensembles with arbitrary means and de Finetti band ensembles. The method of moment was used to obtain the almost sure limit of the appropriately rescaled operator norms for centered Wigner ensembles; then, let M ∈ Mat_nxn_(C) ser una be a matrix. The norm of matrix of operators of M is defined as:

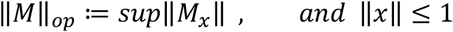

In that case x ∈ C^n^ y ||. || is a normalized vector of C^n^.

#### Teorem 1. Bai-Yin Law

The limiting spectral distribution for matrix Wigner W_n_ is the upper limit of Bai-Yin [25]:

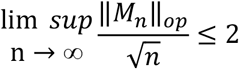

As result, the normalized version is defined like 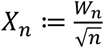

#### Teorem 2. Semicircle for Wigner Distribution

If the W_n_ (*W*_*n*_)_*n*≥1_ is a sequence of Wigner matrices, let µn be the probability measure [9]:

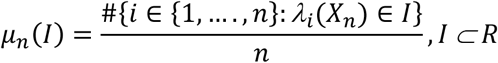

In this case the *λ*_*n*_ (*X*_*n*_) ≤ ⋯.. ≤ *λ*_*n*_ (*X*_*n*_) *∈ R* are the eigenvalues (eigenvalues) of X_n_. As consequence, the µ_n_ weakly converges to the semicircle distribution:

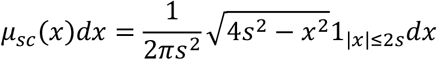

### 2.2 Definitions of the Method of Moments for Probability Measures

One of the most iconic and straightforward tests of the Wigner macroscopic random matrix scale is that it uses the method of the *moment*. This approach is based on the intuition that the eigenvalues of the Wigner matrices are distributed according to a limiting law, which is the semicircle distribution µ_sc_. The moments of the empirical distribution µ_n_ correspond to sampling moments of the limit distribution, where the number of samples is given by the size of the matrix [26].

To calculate the k-th moment with the µ-law of a random variable of X, which is the expectation of the E (X^k^), the eigenvalues of X_n_ are denoted by *λ*_j_(X_n_) with order of *λ*_1_ (*X*_*n*_) ≤ *λ*_2_ (*X*_*n*_) … .. ≤ *λ*_*n*_ (*X*_*n*_). Note that X_n_ can be diagonalized since it is Hermitian matrix. In fact, it has *X*_*n*_ *= U*^*t*^ *D*_*n*_ *U* where *D*_*n*_ = *diag*(*λ*_1_ (*X*_*n*_), *λ*_2_ (*X*_*n*_), … ., *λ*_*n*_ (*X*_*n*_)) Therefore, they are obtained for the k-th moment [13]:

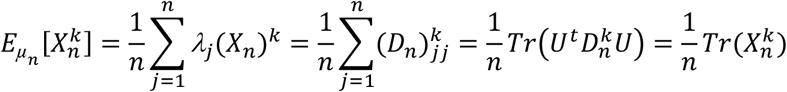

This is a very useful method, in particular considering it does not make any assumptions on the target to measure *µ*_n_. In the literature on random matrices, this condition is often used the method of moments, see [26]. In summary, this is the theorem that is used when applying the method of moments to random matrix theory.

The empirical spectral distributions of random matrices, whose are K-valued in their entries have absolute moments of all orders. Then if where (m_k_)_k_ is a sequence of real numbers that satisfy the Carleman condition, then (σ_n_)_n_ converges in expectation to a probability measure µ on (R, B) with moments (m_k_)_k_. The k-th random moment is given by which is a real-valued random variable whose expectation is finite.

The moments of the semicircle law are given by [13]:

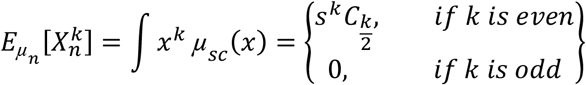

Where Cn is the n-th Catalan number, which is given directly of the binomial coefficients, given by [13]:

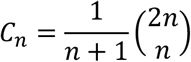

The catalan numbers are elements of the sequence of natural numbers. As a result of the semicircle law, it is the unique distribution where the k-th moments that are given by the Calatan numbers [26]:

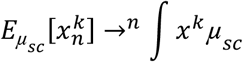

Consequently:

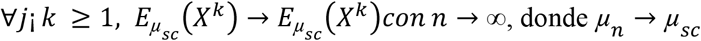

But the Catalan numbers are not only the (even) moments of the semicircle distribution. They also appear as the solution to various combinatorial problems, see [27] or [28], for example.

The method of moments for random probability works as follows: If one wants to show weak convergence of random probability measures in expectation, in probability or almost surely, it will suffice to show that the random moments converge in expectation, in probability or almost surely.

### 2.3 Implementation and Data

#### 2.3.1 COVID-19 Data Analysis

This section presents a COVID-19 risk analysis for the regions of Mexico. The data used here are open data, and it can be found on the website of the federal government in Mexico in the section of the Secretariat of Epidemiology^1^.

The Mexico COVID-19 database has the following hierarchical variables (see Figure 1): 1) positive patients and negative patients, 2) symptomatic patients, and 3) hospitalized and non-hospitalized patients.

**Figure 1.**
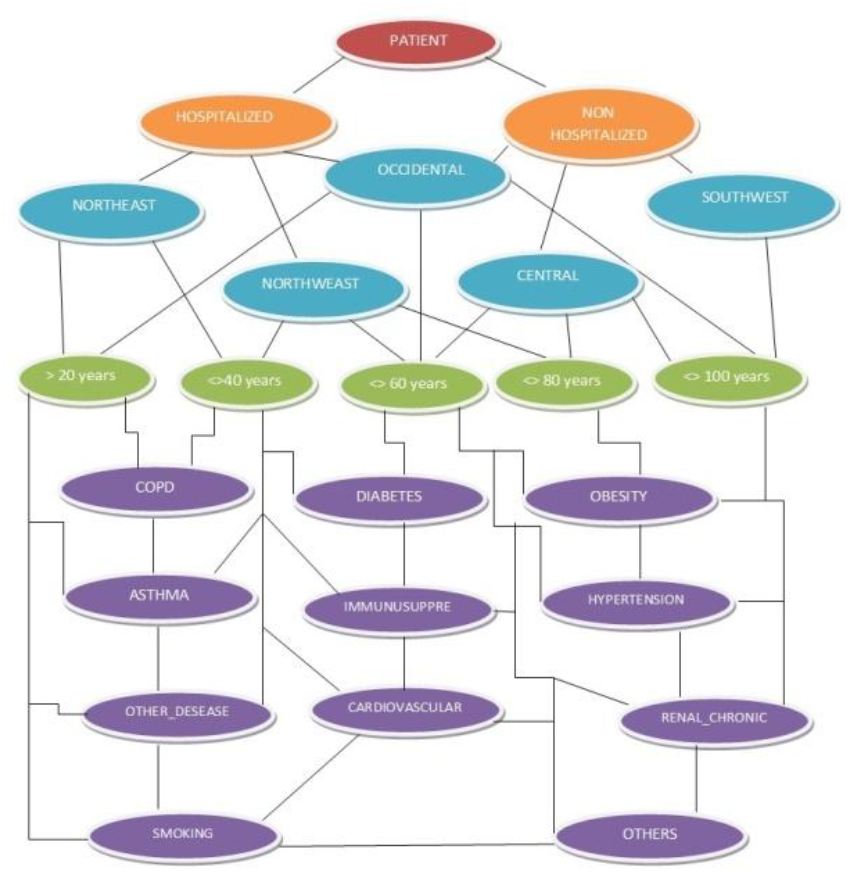
Network database was constructed with hierarchical variables identified by 5 different colors as follows: maroon, orange, blue, green, and purple, figure extracted from Nolasco-Jáuregui O., et al. 2021 [30].

The geographical information of the infected patients by the virus in Mexico is divided into 5 regions (see them in blue color in Figure 1). Let us remember that Mexico has 32 federal states, each with a particular political, economic, population and social situation. The federal states are grouped by regions as: Northwest Region (R1), Northeast Region (R2), West Region (R3), Central Region (R4) and Southeast Region (R5). The R1 has the following states: Baja California, Baja California Sur, Chihuahua, Sinaloa and Sonora. In R2 are: Coahuila, Durango, Nuevo León, San Luis Potosí and Tamaulipas. For the R3 there are: Mexico City, State of Mexico, Guerrero, Hidalgo, Morelos, Puebla and Tlaxcala. In R4 have: Aguascalientes, Colima, Guanajuato, Jalisco, Michoacán, Nayarit, Querétaro and Zacatecas. And then, R5 covers the states of Campeche, Chiapas, Oxaca, Quintana Roo, Tabasco, Veracruz, and Yucatán.

Symptomatic patients are characterized by presenting the major COVID-19 symptoms, these cases show symptoms such as cough, sore throat, fever, or shortness of breath. Once the viral detection test has been applied, if the patients are tested positive they were classified as positive patients and assume to have the virus, otherwise, they were consider as negative patients.

At the first position on the hierarchical variables are positive patients (see them in maroon color in Figure 1); for these cases are following subsections: the symptom onset date, clinic admission date and clinic exit date. At the second place on the hierarchical variables are symptomatic patients, whose are subsectioned as hospitalized patients and non-hospitalized patients (see them in orange color in Figure 1). For symptomatic patients with severe to critical disease or those who are severely immunocompromised, the health experts admitted them at the clinic immediately and were classified as hospitalized patients in our database. For symptomatic patients with mild to moderate disease and not severely immunocompromised, the health experts recommended that they must keep a strict quarantine at home and were classified as non-hospitalized patients in our database.

The study project report is based on a comparison of the hospitalized and non-hospitalized patients with the comorbidities of the patients and their exposure risk to the virus in different regions of Mexico. In these statistical analyses, the principal comorbidities (see them in purple color in Figure 1) on patients are detailed, such as diabetes (D), COPD (CO), asthma(A), Immunosuppression (IM), hypertension (HY), cardiovascular (CA) problems [29], chronic kidney disease (RE), obesity (OB), and others diseases (OD) [31]. People suffering from any comorbidities are at increased risk of severe COVID-19 infection [32-33], the diseases mentioned above play an important role in the possible recovery of patients who have acquired the virus [34-35]. Figure 1 describes the Mexico COVID-19 database extraction and their hierarchical variables. It should be emphasized that the period of analysis of the database corresponds from April 12 to October 5, 2020 (761 665 Patients), giving a total of 176 daily record files.

## 3-. Results

In this proposal we apply the theory of the random matrix in the probability measure that appears as the limit of the empirical spectral distribution as characteristically by the Wigner semicircle law of the hierarquical network shows in Figure 1 for every region of Mexico by comorbidities for hospitalized (H) patients and ambulatory (N) patients, see Figure 2. The Figure 2 is a beautiful graphical representation of the Wigner semicircles overlapping as a result of this propose where it is easy for the readers obtain conclusions about the comorbidity of the regions of Mexico and their COVID-19 patients.

**Figure 2.**
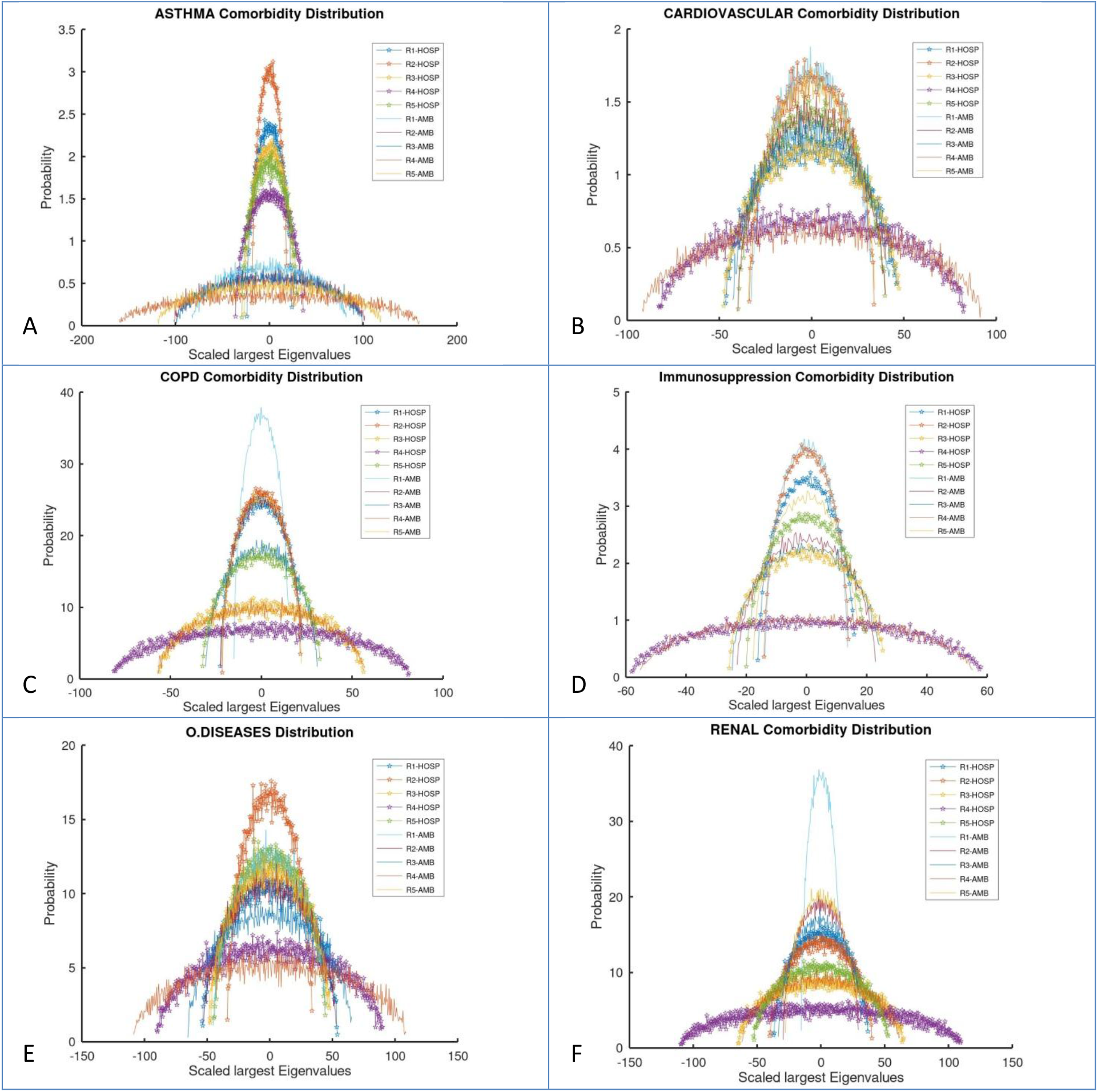
Wigner semicircles overlapping by comorbidities.

In Figure 2-A of asthma comorbidity, it can be observed that R2 presents a greater probability with this morbidity in the case of hospitalized patients; while there is a lower probability in ambulatory patient cases. Consequently, the radius of R2 indicates that the number of hospitalized patients is fewer than ambulatory patient cases. Following up to R2 is the R1, the R1 also has highest cases in hospitalized patients with this comorbidity. But R1 is superior than R2 in ambulatory cases. The radius in outpatients is more platykurtic than in hospitalized ones, since the distribution it is more leptokurtic, see Table 1.

**Table 1.**
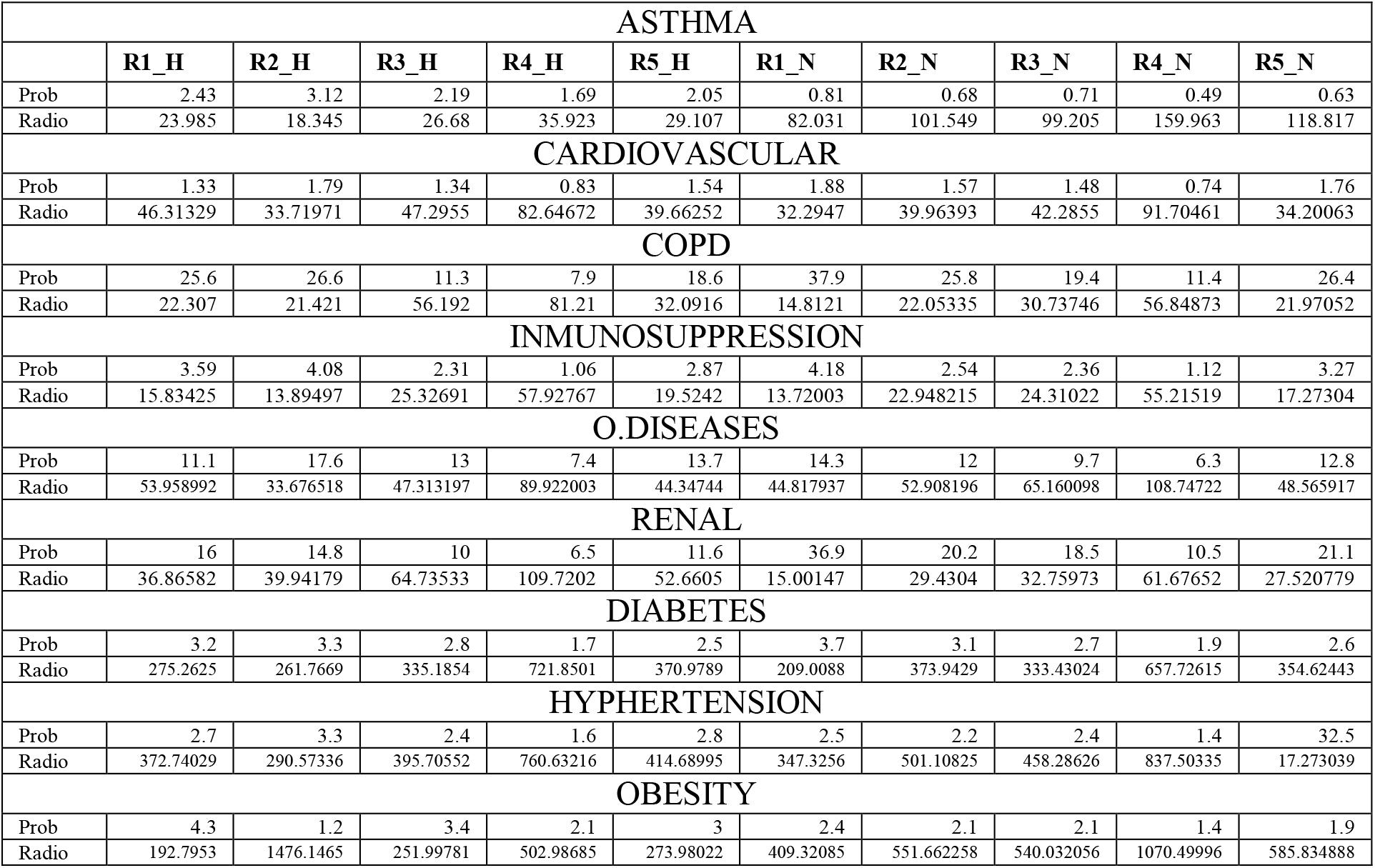
Maximum values of probability and radius of the Wigner semicircles, of the 9 comorbidities reported in Mexico.

| ASTHMA |  |  |  |  |  |  |  |  |  |  |
| --- | --- | --- | --- | --- | --- | --- | --- | --- | --- | --- |
|  | R1_H | R2_H | R3_H | R4_H | R5_H | R1_N | R2_N | R3_N | R4_N | R5_N |
| Prob | 2.43 | 3.12 | 2.19 | 1.69 | 2.05 | 0.81 | 0.68 | 0.71 | 0.49 | 0.63 |
| Radio | 23.985 | 18.345 | 26.68 | 35.923 | 29.107 | 82.031 | 101.549 | 99.205 | 159.963 | 118.817 |
| CARDIOVASCULAR |  |  |  |  |  |  |  |  |  |  |
| Prob | 1.33 | 1.79 | 1.34 | 0.83 | 1.54 | 1.88 | 1.57 | 1.48 | 0.74 | 1.76 |
| Radio | 46.31329 | 33.71971 | 47.2955 | 82.64672 | 39.66252 | 32.2947 | 39.96393 | 42.2855 | 91.70461 | 34.20063 |
| COPD |  |  |  |  |  |  |  |  |  |  |
| Prob | 25.6 | 26.6 | 11.3 | 7.9 | 18.6 | 37.9 | 25.8 | 19.4 | 11.4 | 26.4 |
| Radio | 22.307 | 21.421 | 56.192 | 81.21 | 32.0916 | 14.8121 | 22.05335 | 30.73746 | 56.84873 | 21.97052 |
| INMUNOSUPPRESSION |  |  |  |  |  |  |  |  |  |  |
| Prob | 3.59 | 4.08 | 2.31 | 1.06 | 2.87 | 4.18 | 2.54 | 2.36 | 1.12 | 3.27 |
| Radio | 15.83425 | 13.89497 | 25.32691 | 57.92767 | 19.5242 | 13.72003 | 22.948215 | 24.31022 | 55.21519 | 17.27304 |
| O.DISEASES |  |  |  |  |  |  |  |  |  |  |
| Prob | 11.1 | 17.6 | 13 | 7.4 | 13.7 | 14.3 | 12 | 9.7 | 6.3 | 12.8 |
| Radio | 53.958992 | 33.676518 | 47.313197 | 89.922003 | 44.34744 | 44.817937 | 52.908196 | 65.160098 | 108.74722 | 48.565917 |
| RENAL |  |  |  |  |  |  |  |  |  |  |
| Prob | 16 | 14.8 | 10 | 6.5 | 11.6 | 36.9 | 20.2 | 18.5 | 10.5 | 21.1 |
| Radio | 36.86582 | 39.94179 | 64.73533 | 109.7202 | 52.6605 | 15.00147 | 29.4304 | 32.75973 | 61.67652 | 27.520779 |
| DIABETES |  |  |  |  |  |  |  |  |  |  |
| Prob | 3.2 | 3.3 | 2.8 | 1.7 | 2.5 | 3.7 | 3.1 | 2.7 | 1.9 | 2.6 |
| Radio | 275.2625 | 261.7669 | 335.1854 | 721.8501 | 370.9789 | 209.0088 | 373.9429 | 333.43024 | 657.72615 | 354.62443 |
| HYPHERTENSION |  |  |  |  |  |  |  |  |  |  |
| Prob | 2.7 | 3.3 | 2.4 | 1.6 | 2.8 | 2.5 | 2.2 | 2.4 | 1.4 | 32.5 |
| Radio | 372.74029 | 290.57336 | 395.70552 | 760.63216 | 414.68995 | 347.3256 | 501.10825 | 458.28626 | 837.50335 | 17.273039 |
| OBESITY |  |  |  |  |  |  |  |  |  |  |
| Prob | 4.3 | 1.2 | 3.4 | 2.1 | 3 | 2.4 | 2.1 | 2.1 | 1.4 | 1.9 |
| Radio | 192.7953 | 1476.1465 | 251.99781 | 502.98685 | 273.98022 | 409.32085 | 551.662258 | 540.032056 | 1070.49996 | 585.834888 |

The Table 1 has the maximum limit value of the Wigner distribution, with this value the reader can takes the probability and even the radius. Note: It is important to mention that the X axis has a scaled largest eigenvalues because is a semicircle, so it has negative and positive values; the radius indicates the absolute value of eigenvalues.

The Figure 2-B shows the cardiovascular comorbidity, it is notable that in almost all regions and type of patients have the same numbers because the radius has similar values; is the R4 that has the same radius even ambulatory and hospitalized patients in this morbidity. In Figure 2-B, it can be observed that R1 presents a greater probability with this morbidity in the case of ambulatory patients; Following up to R1 is the R2, the R2 has highest probability in hospitalized patients with this comorbidity, see Table 1.

The Figure 2-C shows the COPD comorbidity, it is notable that in this morbidity the probability value is highest then the last ones diseases in all cases. In this comorbidity, the highest radius is of the R4 in hospitalized patients, as result this region of Mexico has the major number of infected with this morbidity. In Figure 2-C, it can be observed that R1 presents the greater probability with this morbidity in the case of ambulatory patients; Following up to R1 is the R2, the R2 has highest probability in both types of patients ambulatories and hospitalized; for both cases the distribution is leptokurtic.

In Figure 2-D of Immunosuppression comorbidity, it can be observed that R1 presents a greater probability with this morbidity in the case of non-hospitalized patients. Following up to R1 is the R2, the R2 also has highest cases in hospitalized patients with this comorbidity. It is notable that the highest radius is of the R4 in both types of patients hospitalized and ambulatories, as result this region of Mexico has the major number of infected with this morbidity.

In Figure 2-E of other diseases, it can be observed that R2 presents a greater probability with these comorbidities in the case of non-hospitalized patients. Following up to R2 is the R1, the R1 also has highest cases in non-hospitalized patients with this comorbidity. It is notable that the highest radius is of the R4 in ambulatory patients, as result this region of Mexico has the major number of infected with this morbidity.

In Figure 2-F of renal diseases, it can be observed that R1 presents a greater probability with these comorbidities in the case of non-hospitalized patients. Following up to R1 is the R2, the R2 also has highest cases in non-hospitalized patients with this comorbidity. It is notable that the highest radius is of the R4 in both types of patients for ambulatories and hospitalized patients, as result this region of Mexico has the major number of infected with this morbidity.

This document has a medullar analysis with a SuperHeat map, see Figure 3. This SuperHeat map as a complement to the previous analysis, the SuperHeat map [36] is used as a correlation analysis between comorbidities of the regions of Mexico and its daily behavior pandemic analysis. The interpretation of the Figure 3 is as following: it has on its left axis the dendrogram [37] that indicates in its farthest lines the representation of the higher hierarchy and towards the center of the map, the lower hierarchies relationship between all elements.

**Figure 3.**
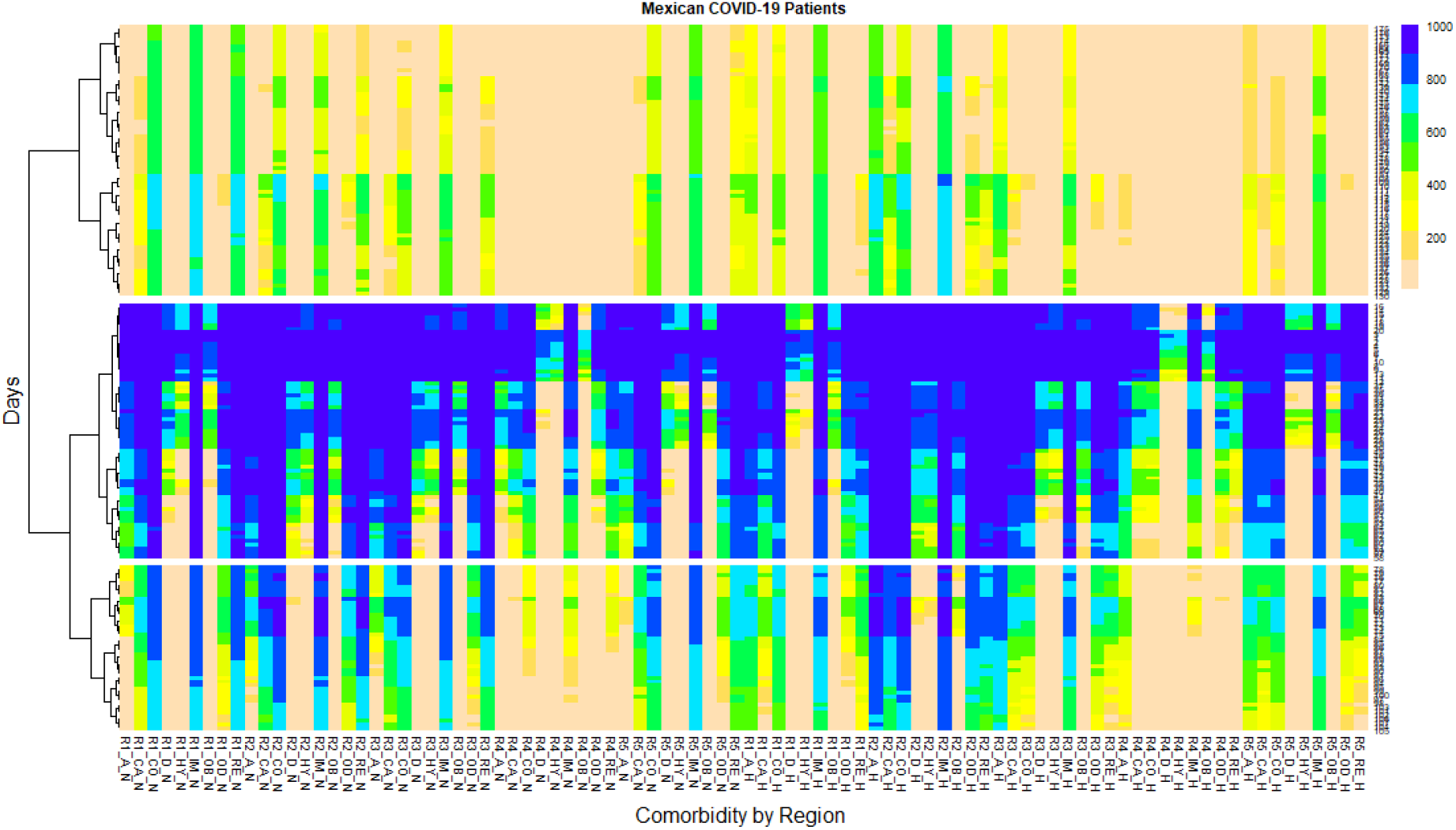
SuperHeat map that shows daily behavior of the pandemic in all regions of Mexico.

In the SuperHeat map you can see 3 large groups, resulting from a Machine Learning algorithm called K-means [36]. It is a type of algorithm clustering that is characterized by indicating how similar they are, that is, the similarity coefficient. The similarity coefficient is achieved with a distance called Euclidean Distance [38]. In Figure 3, the right axis indicates the number of days with a pandemic (from April 12 2019 to October 4, 2020) and its density of infected cases for 176 days. The color scale on the right side of the map indicates the density of infected ranging from 1000 in blue to 200 in pink color corresponding to daily cases.

It is notable that the SuperHeat has three large groupings or clustering. The first clustering has the lowest scale of infections around 200 (75 to 176 day with pandemic). The second clustering has the highest density of infections occurred in the first days of the pandemic since day 1 to day 50, practically all regions and in all comorbidities the patients N and H were infected, and there were about 1000 daily cases of contagion (blue color), see Figure 3. The third clustering was result of the selection of the days that have the highest number of transitions of infections ranging from 1000 to less than 200 daily infections in all regions and by the type of comorbidity of the patients. The third clustering is around days 51 to 74 of pandemic in Mexico, see Figure 3.

In the first clustering analysis, there are 4 cases that have an average prolongation of density of N infected patients for a longer time, alternating between 800 and 600 daily infected cases from day 75 to 176 with the pandemic; where R1 in patients with COPD, Renal and Immunosuppression comorbidities of ambulatory patients and the R5 with Immunosuppression of ambulatory patients too. There are only 3 cases that have a prolongation of average density of infected patients for more than 800 and 600 daily cases of H patients: R1 in patients with Immunosuppression and R2 with Asthma and Immunosuppression. The rest of the regions and comorbidities among the patients hospitalized and non-hospitalized are found alternating with a density of infected around of 400 and less than 200 daily cases. It is very important to note that in this clustering has reported the least number of cases of H, the majority of infected are N and they are in house for their recovery.

In the second clustering analysis, in all regions both N and H patients, they have the highest density of infections with around 1000 daily cases. For non-hospitalized patients, R1 in patients with COPD, Immunosuppression, Renal have the prolongation of the average density of infected patients for the longest time ranging from 1000 to 800 daily of almost 50 days of pandemic. The R2 in patients with Cardiovascular, COPD, Renal, Other Diseases and Immunosuppression with NH have the prolongation of the average density of infected patients for a longer time ranging from 1000 to 800 daily of almost 50 days of pandemic. R3 in patients with Cardiovascular, COPD, Renal, Immunosuppression with NH have the average density prolongation of patients infected for longer periods ranging from 1000 to 800 daily of almost 50 days of pandemic. R5 in patients with Cardiovascular, COPD, Immunosuppression and Renal with NH have the prolongation of the average density of infected patients for a longer time ranging from 1000 to 800 daily of almost 50 days of pandemic. For Hospitalized patients, R2 is the only region that has the prolongation of mean density of infected patients by greater time ranging from 1000 to 800 daily of almost 50 days of pandemic, in comorbidities such as Asthma, Cardiovascular, COPD and Immunosuppression.

## Conclusions

At the end of 2019, humanity faced the challenge of a new pandemic called COVID-19. This disease has claimed the lives of thousands of people, mainly in developing economies. Therefore, this research presents a regional analysis of COVID-19 in Mexico. The analysis period is from April 12 to October 5, 2020 (761,665 patients). From Random Matrix Theory, moments Wigner’s law of the semicircle and Machine Learning in SuperHeat maps, the probability distribution of 9 comorbidities in Mexican patients for 5 geographic regions was study.

The COPD comorbidity, renal diseases and other diseases represent the highest probability in infected patients who have been hospitalized (H). In the case of COPD, renal diseases and other diseases in region 1 and region 2 present the highest probability value in hospitalizations. On the other hand, these same comorbidities are also found in ambulatory patients, where high levels of probability prevail among those infected.

Undoubtedly this methodology is unique and it is applicable to COVID-19 data for many countries in the world in comorbidities of patients. In general the RMTs can work to analyze the multivariate behavior of large data as it is done in this work and obtain fast visual conclusion about the eigenvalues and their probability, see Figure 2. This Figure 2 is a gorgeous graphical representation of the Wigner semicircles overlapping as a result of this propose where it is easy for the readers to obtain conclusions about the comorbidity of the regions of Mexico and their COVID-19 patients.

Figure 3 concludes the daily analysis. The SuperHeat map is a complex graph that includes the correlation analysis. This is used to study the comorbidities of the regions of Mexico and its daily analysis of the pandemic behavior.

## Data Availability

https://www.gob.mx/cms/uploads/attachment/file/604001/Datos_abiertos_hist_ricos_2020.pdf

https://github.com/OraliaNJ/COVID-19_Mex_Analysis

## Abbreviations

(D): Diabetes
(CO): COPD
(A): Asthma
(IM): Immunosuppression
(HY): Hypertension
(CA): Cardiovascular
(RE): chronic kidney disease
(OB): obesity
(OD): others diseases
(R1): Region 1
(R2): Region 2
(R3): Region 3
(R4): Region 4
(R5): Region 5
(N): Non-hospitalized patient
(H): Hospitalized patients

## Footnotes

1 **Open Data of the Ministry of Health in the Department of Epidemiology from the Government of Mexico**. Historical bases COVID-19. Available on: https://www.gob.mx/cms/uploads/attachment/file/604001/Datos_abiertos_hist_ricos_2020.pdf. Our file repository analysis are available on: https://github.com/OraliaNJ/COVID-19_Mex_Analysis

## References

1. Hui, D. S., Azhar, E. I., Madani, T. A., Ntoumi, F., Kock, R., Dar, O., … & Petersen, E. (2020). The continuing 2019-nCoV epidemic threat of novel coronaviruses to global health—The latest 2019 novel coronavirus outbreak in Wuhan, China. International journal of infectious diseases, 91, 264–266.

2. Aragón-Nogales, R., Vargas-Almanza, I., & Miranda-Novales, M. G. (2019). COVID-19 por SARS-CoV-2: la nueva emergencia de salud. Revista mexicana de pediatría, 86(6), 213–218.

3. WHO. Coronavirus. Retrieved July 3, 2021. From the World Health Organization. Available on: https://www.who.int/health-topics/coronavirus#tab=tab_1.

4. Girko, V. L. (1985). Spectral theory of random matrices. Russian Mathematical Surveys, 40(1), 77–120.

5. Melo, M. (2015). Applications of Random Matrices to Image Processing for Image Denoising. Available on: https://repository.lib.fit.edu/handle/11141/736

6. Bai, Z. D. (1997). Circular law. The Annals of Probability, Vol. 25, No. 1, pp. 494–529. Available on: https://projecteuclid.org/euclid.aop/1024404298#:∼:text=It%20was%20conjectured%20in%20the,is%20called%20the%20circular%20law

7. Namaki, A., Ardalankia, J., Raei, R., Hedayatifar, L., Hosseiny, A., Haven, E., & Jafari, R. (2020). Analysis of the Global Banking Network by Random Matrix Theory. arXiv preprint 2007.14447.

8. Kirsch, W., & Kriecherbauer, T. (2020). Random matrices with exchangeable entries. Reviews in Mathematical Physics, 32(07), 2050022.

9. Arous, G. B., & Guionnet, A. (2011). Wigner matrices. In The Oxford Handbook of Random Matrix Theory (pp. 433–451). Oxford University Press. Available on: http://perso.ens-lyon.fr/aguionne/RMTChap21.pdf

10. Dos Santos, F. C., Federspiel, S., & Schammo, A. (2018). Spectral Theory of Random Matrices. University of Luxembourg. Retrieved on November 20th, 2020 from Department of Mathematics: https://math.uni.lu/eml/projects/reports/random-matrices.pdf

11. Hirviniemi, O. (2017). Fundamental properties of random Hermitian matrices, pp. 29– Available on: https://helda.helsinki.fi/handle/10138/176233

12. O’Rourke, S. (2012). A note on the Marchenko-Pastur law for a class of random matrices with dependent entries. Electronic Communications in Probability, Vol. 17. Available on: https://projecteuclid.org/euclid.ecp/1465263161

13. Valkó, Benedek. Lecture 1: Basic random matrix models (2009). Department of Mathematics. University of Wisconsin. Available on: http://www.math.wisc.edu/∼valko/courses/833/2009f/lec_01.pdf

14. Fleermann, M. (2019). Global and Local Semicircle Laws for Random Matrices with Correlated Entries (Doctoral dissertation, FernUniversität in Hagen).

15. Allez, R. (2012). Chaos multiplicatif Gaussien, Matrices aléatoires et applications (Doctoral dissertation, Paris 9).

16. Masucci, A. M. (2011). Moments method for random matrices with applications to wireless communication (Doctoral dissertation, Supélec).

17. Benaych-Georges, F., & Knowles, A. (2016). Lectures on the local semicircle law for Wigner matrices. arXiv preprint 1601.04055.

18. Medel-Ramírez, C., & Medel-Lopez, H. (2020). Data Mining for the Study of the Epidemic (SARS-CoV-2) COVID-19: Algorithm for the Identification of Patients (SARS-CoV-2) COVID 19 in Mexico. Available at SSRN 3619549.

19. Parra-Bracamonte, G. M., Lopez-Villalobos, N., & Parra-Bracamonte, F. E. (2020). Clinical characteristics and risk factors for mortality of patients with COVID-19 in a large data set from Mexico. Annals of epidemiology, 52, 93–98.

20. Najera, H., & Ortega-Avila, A. G. (2021). Health and Institutional Risk Factors of COVID-19 Mortality in Mexico, 2020. American journal of preventive medicine, 60(4), 471–477.

21. Prieto, K., Chavez-Hernandez, M., & Romero-Leiton, J. P. (2021). On mobility trends analysis of COVID-19 dissemination in Mexico City. medRxiv.

22. Porikli, F., Tuzel, O., & Meer, P. (2006, June). Covariance tracking using model update based on lie algebra. In 2006 IEEE Computer Society Conference on Computer Vision and Pattern Recognition (CVPR’06) (Vol. 1, pp. 728-735). IEEE.

23. Tracy, C. A., & Widom, H. (2000). The distribution of the largest eigenvalue in the Gaussian ensembles: β= 1, 2, 4. In Calogero—Moser—Sutherland Models (pp. 461–472). Springer, New York, NY.

24. Bogachev, L., Molchanov, S. A., & Pastur, L. A. (1992). On the density of states of random band matrices.(Russian) Mat. Zametki 50 (1991), no. 6, 31–42; translation in Math. Notes, 50, 5–6.

25. Bai, Z. D., & Yin, Y. Q. (1988). Necessary and sufficient conditions for almost sure convergence of the largest eigenvalue of a Wigner matrix. The Annals of Probability, 1729–1741.

26. Anderson, G. W., Guionnet, A., & Zeitouni, O. (2010). An introduction to random matrices (No. 118). Cambridge university press.

27. Bauer, H. (2002). Wahrscheinlichkeitstheorie. Walter de Gruyter.

28. Charalambides, C. A. (2018). Enumerative combinatorics. CRC Press.

29. Jordan, R. E., Adab, P., & Cheng, K. (2020). Covid-19: risk factors for severe disease and death.

30. Nolasco-Jauregui, O., Quezada-Tellez, L. A., Rodriguez-Torres, E. E., & Fernandez-Anaya, G. (2021). COVID-19 Patients Analysis using Superheat Map and Bayesian Network to identify Comorbidities Correlations under Different Scenarios. medRxiv.

31. Fang, L., Karakiulakis, G., & Roth, M. (2020). Are patients with hypertension and diabetes mellitus at increased risk for COVID-19 infection?. The Lancet. Respiratory Medicine, 8(4), e21.

32. Phelps, M., Christensen, D. M., Gerds, T., Fosbøl, E., Torp-Pedersen, C., Schou, M., … & Gislason, G. (2021). Cardiovascular comorbidities as predictors for severe COVID-19 infection or death. European Heart Journal -Quality of Care and Clinical Outcomes, 7(2), 172–180.

33. Mehra, M. R., Desai, S. S., Kuy, S., Henry, T. D., & Patel, A. N. (2020). Cardiovascular disease, drug therapy, and mortality in Covid-19. New England Journal of Medicine, 382(25), e102.

34. Valente-Acosta, B., Hoyo-Ulloa, I., Espinosa-Aguilar, L., Mendoza-Aguilar, R., Garcia-Guerrero, J., Ontanon-Zurita, D., … & Moreno-Sanchez, F. (2020). COVID-19 severe pneumonia in Mexico City-First experience in a Mexican hospital. medRxiv.

35. Kassir, R. (2020). Risk of COVID-19 for patients with obesity. Obesity Reviews, 21(6).

36. Barter, R. L., & Yu, B. (2018). Superheat: An R package for creating beautiful and extendable heatmaps for visualizing complex data. Journal of Computational and Graphical Statistics, vol. 27, no 4, p. 910–922.

37. Wu, L., Peng, Y., Fan, J., Wang, Y., & Huang, G. (2021). A novel kernel extreme learning machine model coupled with K-means clustering and firefly algorithm for estimating monthly reference evapotranspiration in parallel computation. Agricultural Water Management, 245, 106624.

38. Lange, M., Zühlke, D., Holz, O., Villmann, T., & Mittweida, S. G. (2014, April). Applications of lp-Norms and their Smooth Approximations for Gradient Based Learning Vector Quantization. In ESANN (pp. 271–276).

